# Ribosome Phenotypes Enable Rapid Antibiotic Susceptibility Testing in *Escherichia coli*

**DOI:** 10.1101/2024.06.18.24309111

**Authors:** Alison Farrar, Piers Turner, Hafez El Sayyed, Conor Feehily, Stelios Chatzimichail, Derrick Crook, Monique Andersson, Sarah Oakley, Lucinda Barrett, Christoffer Nellåker, Nicole Stoesser, Achillefs Kapanidis

## Abstract

Rapid antibiotic susceptibility tests (ASTs) are an increasingly important part of clinical care as antimicrobial resistance (AMR) becomes more common in bacterial infections. Here, we use the spatial distribution of fluorescently labelled ribosomes to detect intracellular changes associated with antibiotic susceptibility in single *E. coli* cells using a convolutional neural network (CNN). By using ribosome-targeting probes, a single fluorescence cell image provides data for cell segmentation and susceptibility phenotyping. Using 50,722 images of cells from an antibiotic-susceptible laboratory strain of *E. coli*, we showed that antibiotics with different mechanisms of action result in distinct ribosome phenotypes, which can be identified by a CNN with high accuracy (99%, 96%, and 91% for ciprofloxacin, gentamicin, and chloramphenicol). With 6 *E. coli* strains isolated from bloodstream infections, we used 34,205 images of ribosome phenotypes to train a CNN that could classify susceptible cells with 92% accuracy and resistant cells with 99% accuracy. Such accuracies correspond to the ability to differentiate susceptible and resistant samples with 99% confidence with just 2 cells, meaning that this method could eliminate lengthy sample culturing steps and could determine in vitro susceptibility with 30 minutes of antibiotic treatment. Our ribosome phenotype method should also be able to identify phenotypes in other strains and species.

## Introduction

Bacterial infections were associated with 14% of all global deaths and the majority of sepsis-related deaths^1^ in 2019. The widespread use of antibiotics in the treatment and prevention of these infections, in medicine and in agriculture, has created a strong evolutionary pressure for microbes resistant to these compounds^2^. In 2019, antimicrobial resistance (AMR) in bacteria caused 1.27 million deaths and was associated with 4.95 million deaths worldwide^3^. Mortality is predicted to rise as high as 10 million deaths per year by 2050 if no action is taken^4^. These challenges motivate the development of new antimicrobial treatments and technologies to mitigate the effects of resistant infections.

Antibiotic susceptibility tests (ASTs) are an essential tool for refining treatment and minimising inappropriate antibiotic use. However, in most clinical microbiology pathways, ASTs are performed after a bacterial pathogen has been cultured and identified, with results available in 12-48 hours for common species^5^. This time delay is often too long to wait in life-threatening infection^6^, leading clinicians to prescribe empirically and use combinations of broad-spectrum antibiotics. In clinical trials, the use of rapid ASTs improves clinical outcomes, decreases the use of broad-spectrum antibiotics, and shortens the time between sample collection and optimal targeted antibiotic treatment^7,8^.

The clinical need for rapid ASTs has motivated the development of new diagnostic technologies to identify the infecting species and characterise susceptibility. Current growth-based ASTs quantify the Minimum Inhibitory Concentration (MIC), a marker of the susceptibility of the isolated organism to an antibiotic^9^, which is typically measured using turbidity. Faster assays based on genotype and cellular morphology are being developed. The bioMerieux BioFire FilmArray system, for example, is a commercial genotype-focused platform utilising multiplex polymerase chain reaction to detect species-specific and resistance-associated genes in syndromic infections (e.g. respiratory, bloodstream, and joint) within an hour^10^. However, PCR methods cannot detect AMR genes that are not present in the PCR probe set, and resistance genes do not always correlate with an isolate’s antibiotic response. A rapid phenotypic test that directly measures the bacterial response may offer advantages over genotypic assays, especially in Gram-negative species which are more likely to have polygenic and combinatorial mechanisms of resistance^5,11^.

Some of the discordances between the genotype and the phenotypic susceptibility may be explained by phenotypic heterogeneity within a bacterial population, leading to phenomena such as persister cells^12,13^. Techniques that directly measure single-cell antibiotic response are advantageous because they can capture this heterogeneity. Many methods have been proposed, including using microscopy to measure growth rate^14,15^, structural changes^16,17^, or cell death^18^; flow cytometry^19^; Raman spectroscopy^20^; and cell impedance^21^. An example of a commercially available phenotypic system is the Accelerate Diagnostics Pheno System, which combines fluorescence in situ hybridization (FISH) for species identification with monitoring of single-cell growth rates to report antibiotic resistance within 7 hours^22^.

Visually apparent changes to the intracellular structure of the bacterial cells can also be used to measure the bacterial antibiotic response. When antibiotics disrupt cellular physiology, long-recognised and characteristic phenotypes develop, which have recently been characterised at scale with high-content imaging^23,24^. Our group showed that such phenotypic effects on the nucleoid and cell membrane can be visualised within 30 minutes and recognised by trained deep-learning models, and that this variability correlates with clinical antibiotic susceptibility^25^. While many novel ASTs have been proposed and developed^5,14–22^, by using single-cell imaging data, we can rapidly and directly capture and assay the diversity of antibiotic response within the cell population.

Here, we present a method for ultra-rapid identification of single-cell antibiotic susceptibility by detecting intracellular changes using ribosome-bound FISH probes (Figure 1a). First, bacteria from the clinical sample are treated with an antibiotic that will induce phenotypic changes in bacteria susceptible to the antibiotic. Following the antibiotic treatment, the cells are fixed, permeabilised, and incubated with species-specific FISH probes. Images of the fluorescent probes are processed and fed to a pre-trained neural network to classify the bacteria as antibiotic-susceptible or antibiotic-resistant. We found that a convolutional neural network (CNN) could learn to recognize the distinct ribosome phenotypes of *E. coli* treated with antibiotics with three different mechanisms of action (ciprofloxacin, a fluoroquinolone, targeting DNA gyrase/topoisomerases; gentamicin, an aminoglycoside, targeting the 30S ribosomal subunit; and chloramphenicol, an amphenicol, targeting the 50S ribosomal subunit). We applied the ciprofloxacin CNN to classify three ciprofloxacin-susceptible and three ciprofloxacin-resistant *E. coli* clinical isolates and found that its accuracy decreased when the isolate’s antibiotic response diverged from the lab strain’s phenotype. Therefore, we developed a CNN trained on images of the clinical isolates, which was able to classify unseen, holdout single-cell images as antibiotic-susceptible with >90% accuracy and antibiotic-resistant with >98% accuracy based on their ribosome phenotypes. Our method advances existing phenotypic ASTs because, when used in combination with multiplexed FISH and species-specific probes, the ribosome fluorescent profile can be used to segment single cells, identify bacterial species, and characterise a cell’s antibiotic response in a single step.

**Figure 1.**
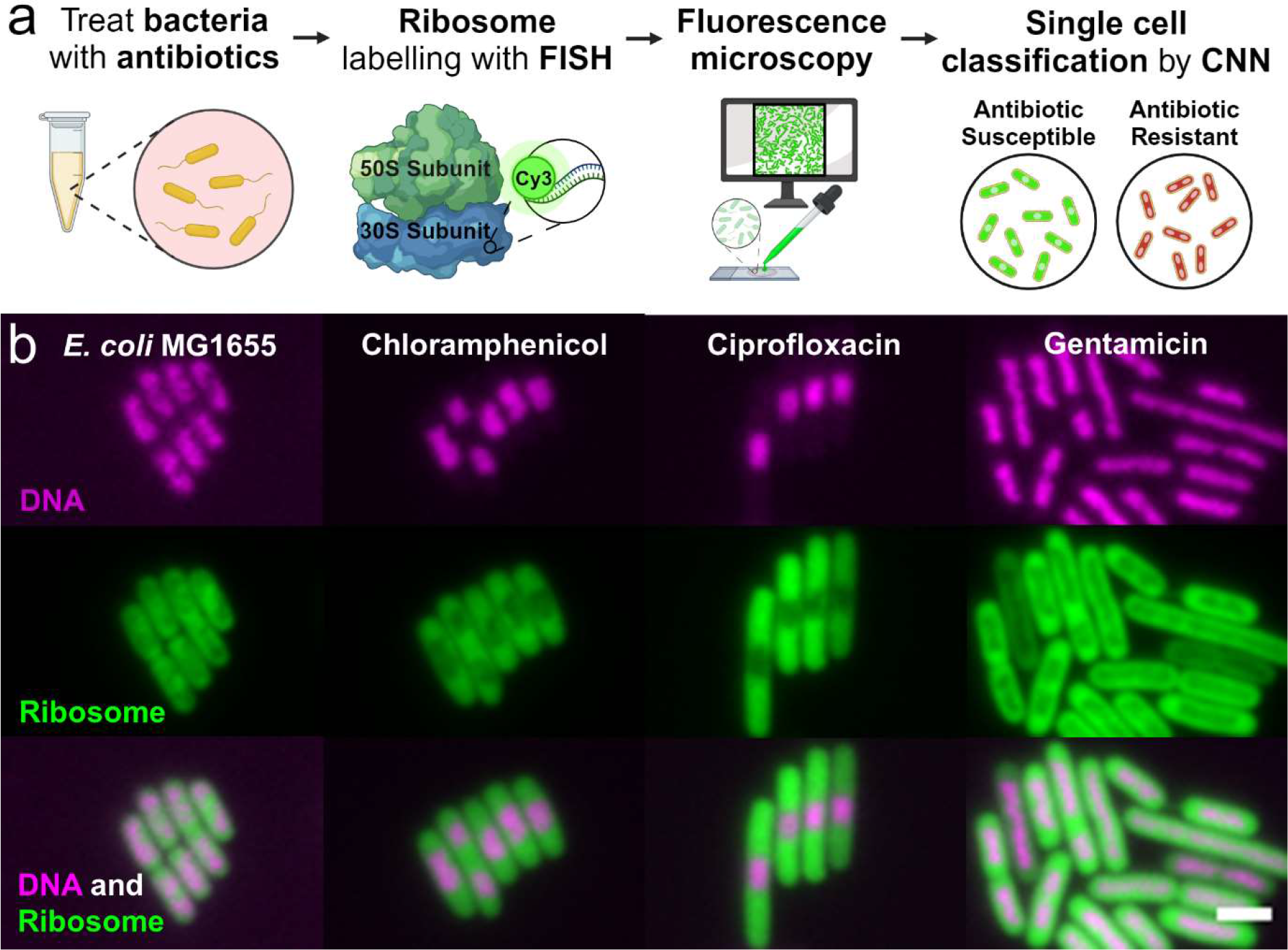
Proposed workflow for using ribosome phenotypes in an ultra-rapid antibiotic susceptibility test. (a) Workflow: First the clinical isolate is treated with antibiotics at a standard concentration for 30 minutes. Then, a standard FISH protocol is used to label the ribosomes with ssDNA fluorescent probes; in this case, EUB338-Cy3 binds a conserved region in the 30S subunit. The samples are imaged on a fluorescence microscope before neural networks use the ribosome signal to segment and then classify the cells as susceptible or resistant to the prescribed antibiotic treatment. (b) Ribosome Phenotypes: Representative fluorescence images are shown of *E. coli* MG1655 with and without antibiotic treatment (magenta, DNA stained with DAPI; green, ribosomes labelled with EUB338-Cy3 probes; combined DNA and ribosome signal). The scale bar is 2 µm. The ribosome density can be seen to anti-correlate with the DNA-dense regions. The untreated panel shows fixed cells with no antibiotic treatment. The chloramphenicol panel shows cells treated with 8 mg/L chloramphenicol (1X EUCAST breakpoint) for 30 minutes before fixation. The ciprofloxacin panel shows the same, treated with 0.5 mg/L ciprofloxacin (1X EUCAST breakpoint). The gentamicin panel shows the same, treated with 40 mg/L gentamicin (20X EUCAST breakpoint).

## Results

### Characterization of the *E. coli* antibiotic response by ribosome subcellular distribution

To train a machine-learning model to classify antibiotic-resistant and antibiotic-susceptible *E. coli*, we characterised the antibiotic-response phenotypes of antibiotic-susceptible cells. Previous work has shown the successful classification of antibiotic-susceptible and resistant *E. coli* by a CNN trained to identify changes in DNA morphology^25^. It has also been shown that DNA-rich and ribosome-rich regions spatially anti-correlate in *E. coli*^26^. Therefore, we reasoned that we may be able to use ribosome phenotypes to classify a bacterium’s antimicrobial response. Because of their space-filling properties, we also hypothesized that the ribosome signal should suffice for both cell segmentation and phenotype analysis, eliminating the need for a membrane dye for cell segmentation. Ribosome fluorescence images may also provide richer spatial and intensity features throughout the cell than images of the nucleoid morphology.

To test our hypothesis, we first characterised the sub-cellular ribosome phenotypes of antibiotic-susceptible *E. coli* MG1655 (Figure 1b). MG1655 is a lab-adapted K-12 derivative that is susceptible to each of the antibiotics used in this work. After treatment with each of the 4 antibiotics individually for 30 minutes, the cells were stained with fluorescent FISH probes to visualize the effects of antibiotic treatment on their internal structure. For this, we used an 18-mer single-strand DNA probe with Cy3 dye conjugated to the EUB338 sequence, which targets a region in the 16S ribosomal RNA conserved in all members of the domain Bacteria^27^.

The antibiotic treatment concentrations were chosen as a multiple of the European Committee on Antimicrobial Susceptibility Testing (EUCAST) breakpoint^9^ for Enterobacterales including *E. coli*, so that an empirical benchmark could be applied to other strains. The EUCAST breakpoint is a defined concentration of antibiotic used to classify a microorganism as antibiotic susceptible and resistant, accounting for clinical factors including antibiotic dosage, target infections, pharmacokinetics, and resistance mechanisms. The MIC of a bacterial isolate can be compared to the EUCAST breakpoint for a given antibiotic and bacterial species to classify it as clinically susceptible (S) or resistant (R).

The biological effect of antibiotic treatment on the susceptible MG1655 bacteria can be clearly seen within 30 minutes. Fluorescence images of the DNA and ribosomes show the characteristic changes in cell spatial organization that occur as the cell responds to the antibiotic (Figure 1b). Comparing the DNA and ribosome signals shows the anti-correlation between DNA and ribosome density within the cell in untreated and antibiotic-treated conditions. The nucleoid compaction caused by chloramphenicol, ciprofloxacin, and gentamicin can be seen as clearly in the ribosome images as in the DNA images. These images also show how the ribosomes fill the cell, allowing the ribosome signal to be used for both cell phenotyping and cell segmentation.

We characterised the ribosome phenotypes from four biological replicates of *E. coli* MG1655 totalling 5,286 untreated cells, 3,215 cells treated with ciprofloxacin (Cip) at 0.5 mg/L (1X EUCAST breakpoint), 5,935 cells treated with gentamicin (Gent) at 40 mg/L (20X EUCAST breakpoint), and 6,439 cells treated with chloramphenicol (Cam) at 8 mg/L (1X EUCAST breakpoint). For *E. coli* MG1655, treatment with chloramphenicol or ciprofloxacin at 1X EUCAST induced phenotypic changes within 30 minutes, but gentamicin treatment concentrations lower than 20X EUCAST did not induce phenotypic changes in most cells in this time frame (Figure S1).

By inspecting the nucleoid and ribosome fluorescence signals along the long axis of the cells, we can further characterise the treatment phenotypes. In untreated *E. coli*, the highest ribosome density was seen in the centre of the cell and in longer cells there were often two ribosome-poor nucleoid regions (Figures 1b; 2a). Ciprofloxacin treatment caused a central, compact nucleoid region (Figures 1b; 2a) and resulted in cells that were longer than untreated cells (Figure 2b). Gentamicin treatment led to a diffuse nucleoid region following the long axis of the cell that was often rod-shaped (Figures 1b; 2a). Chloramphenicol treatment caused nucleoid compaction compared to the untreated phenotype, causing either a centralised DNA region or two dense DNA regions (Figures 1b; 2a). All antibiotic treatments resulted in cells with significantly different in average length and average width compared to the untreated phenotype (Mann-Whitney non-parametric hypothesis test p<0.05) (Figure 2b-c). The antibiotic treatment phenotypes in our images aligned with those found in previous work^28–30^ and with the mechanism of action of each antibiotic (Figure 1b; 2). Because these phenotypes are quantifiable by ribosome fluorescence intensity mapping and identifiable by the human eye, it follows that a neural network could be trained to associate them with an antibiotic treatment response.

**Figure 2.**
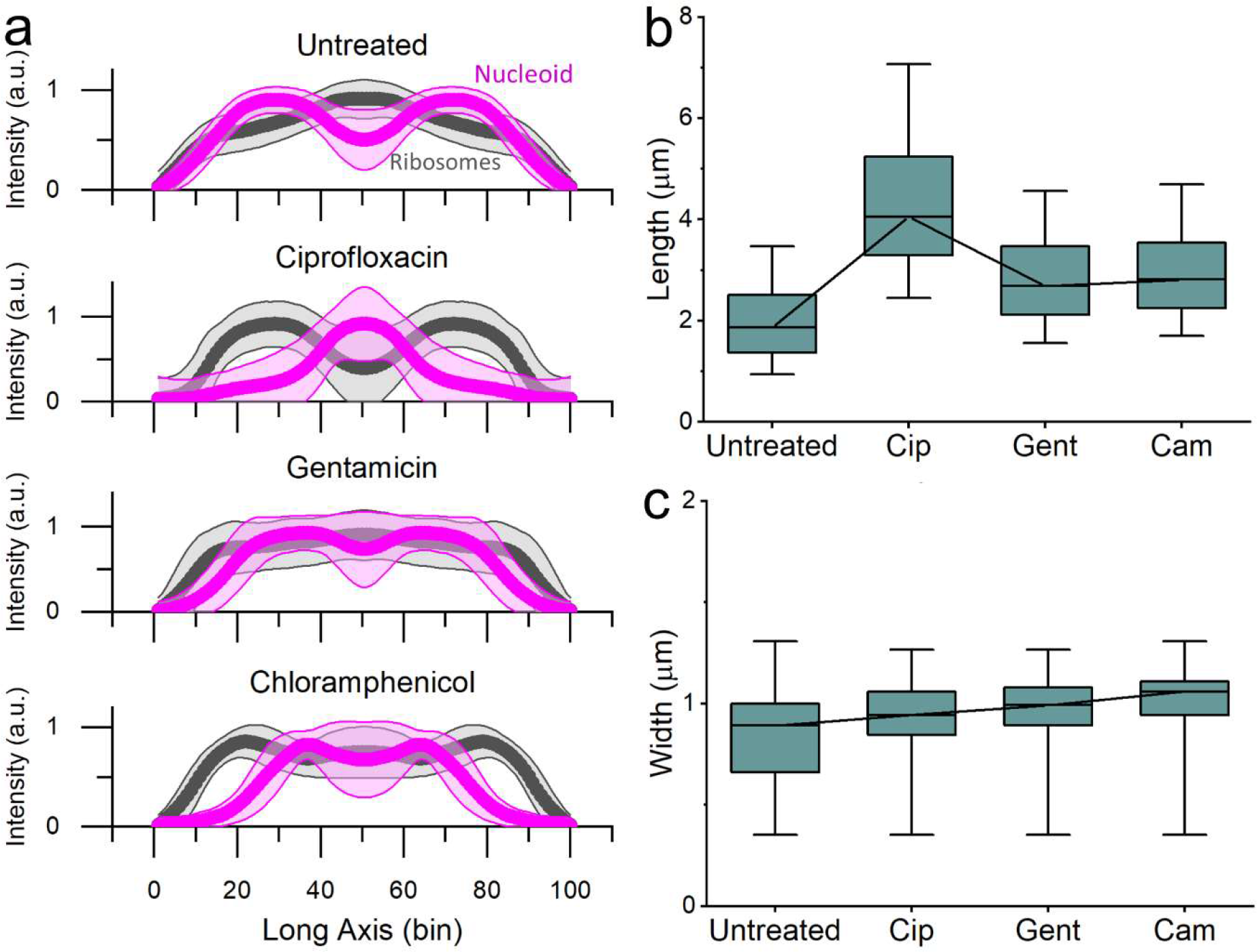
Ribosome intensity line profiles reveal ribosome-nucleoid anti-correlation and characteristic phenotypes of antibiotic response. (a) Along the long axis of the cell, the mean normalised ribosome (Cy3, grey) and nucleoid (DAPI, magenta) intensities are calculated for each of 100 bins. The shading shows ± 1 standard deviation of the mean. The untreated *E. coli* line profiles show two nucleoid-rich regions, correlated with decreased ribosome intensity. This figure is composed of profiles from 5,286 untreated *E. coli* MG1655. The ciprofloxacin panel shows the same, for 3,215 *E. coli* MG1655 treated with 1X EUCAST ciprofloxacin for 30 minutes. The line profile shows a central, compact nucleoid region with greater segregation from the ribosomes. The gentamicin panel shows the same, for 5,935 *E. coli* MG1655 treated with 20X EUCAST gentamicin for 30 minutes. This line profile shows a diffuse nucleoid region along the long axis of the cell with less ribosome-nucleoid segregation. The chloramphenicol panel shows the same, for 6,438 *E. coli* MG1655 treated with 1X EUCAST chloramphenicol. This line profile shows nucleoid compaction compared to the untreated phenotype, with a centralised DNA region or two dense DNA regions. (b) The cell lengths (μm) are shown for untreated *E. coli* MG1655 and for each of the antibiotic treatments. The box shows the 25%∼75% percentile range, the bars show the 1%-99% percentile range, and the line denotes the median. Each antibiotic results in a length distribution statistically different from the untreated population with p<0.05 by the Mann-Whitney non-parametric test. (c) The cell widths (μm) are shown for untreated *E. coli* MG1655 and for each of the antibiotic treatments. The box shows the 25%∼75% percentile range, the bars show the 1%-99% percentile range, and the line denotes the median. Each antibiotic results in a width distribution statistically different from the untreated population with p<0.05 by the Mann-Whitney non-parametric test.

Antibiotic-susceptible ribosome phenotypes are identified accurately by a neural network To train neural networks that can robustly identify the ribosome phenotypes resulting from antibiotic treatments, our fluorescence images were pre-processed prior to their use as training data. First, each single-channel image was segmented by a custom CellPose^31^ model trained to segment *E. coli* by ribosome fluorescence profiles. The segmentations were subsequently curated to refine the outlines and remove cells that were outside of the field of view, overlapping, or outside of the focal plane^32^. To regularise learning and prevent overfitting, each segmentation was used to create a 64x64 zero-filled image with the ribosome fluorescence in the centre, and augmentations (e.g., brightness normalisation, random noise, and geometric transformations) were applied before each image was loaded into the training dataset (Figure S2).

To test the reliability and accuracy of the neural network in differentiating the antibiotic-susceptible phenotypes from untreated *E. coli* MG1655, a rotating holdout test was performed (Figure 3a). For each experiment, a model was independently trained and validated on data from three of the biological replicates and tested on the fourth. The validation and testing datasets were balanced to include an equal number of untreated and antibiotic-treated cell images to minimise prediction biases. The average balanced accuracy of the models on the four test datasets exceeded 90% for all antibiotics: ciprofloxacin∼99±1.6%, gentamicin∼96±7.2%, chloramphenicol∼91±13.8% (Figure 3b). Looking at the models’ predictions, the cells that were classified with high confidence (Figure S5) demonstrated the characteristic antibiotic phenotypes (Figure 2).

**Figure 3.**
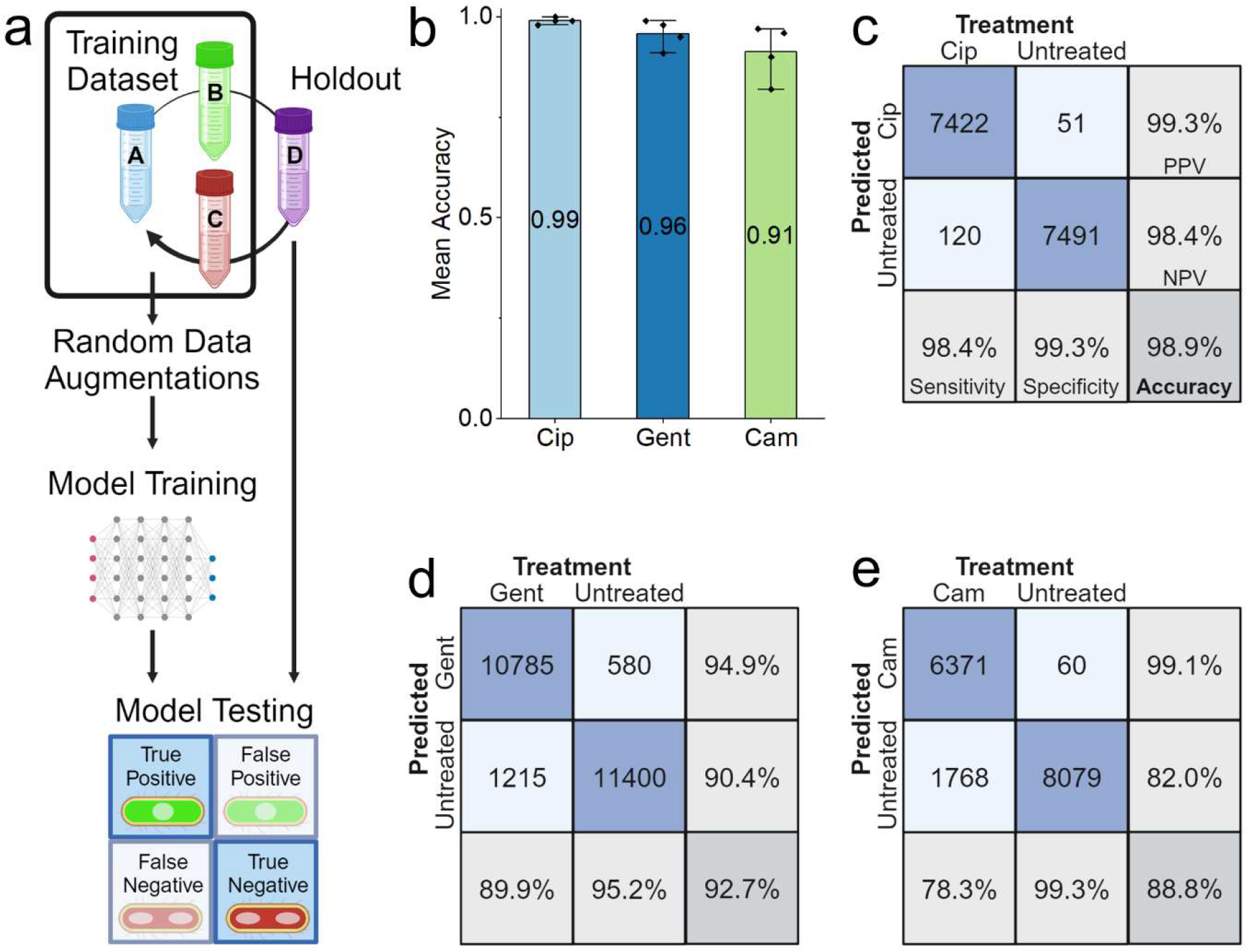
Ribosome phenotype recognition is robust across biological replicates. (a) Four biological replicates of *E. coli* MG1655 were tested for each antibiotic and for the untreated condition. To test phenotype robustness and repeatability, a holdout cross-validation was performed in which each model was trained and validated on images from three of the biological replicates and tested on images from the fourth. The training images received random data augmentations before being passed to the model, whereas the holdout dataset was passed directly to the model for testing. (b) The balanced accuracy of the ribosome phenotype classifier is shown for each antibiotic. Each point represents a biological replicate. The mean balanced accuracy is shown as on each column and the error bars indicate the 95% confidence interval of the mean on the four biological replicates. (c) Confusion matrices for the ribosome phenotype classifier. The total number of cells is a sum of the results from four experiments, each with a model trained on three biological replicates and tested on a fourth holdout replicate. The number of images in each class is shown, along with the percentage of cells for each treatment condition. The treatment condition is shown on the columns and the model’s predicted classification is shown on the columns. Right column: positive predictive value (PPV) and negative predictive value (NPV) of the model’s predictions are shown. Bottom row: accuracy of the model on antibiotic-treated cells (Sensitivity), accuracy of the model on untreated cells (Specificity), and the Balanced Accuracy (Accuracy) are shown. See *Accuracy Metrics* for details.

We then examined the confusion matrices for the four holdout datasets, summed together. The ciprofloxacin phenotype model was the most accurate, with a very high average balanced accuracy of 98.9%, sensitivity of 98.4%, and specificity of 99.3% (Figure 3c). This may be because the ciprofloxacin response causes two phenotypic changes – elongated cells and condensed, central nucleoid – both of which can be used by the model in the classification task (Figure S3). The gentamicin phenotype model was also highly accurate, achieving an average balanced accuracy of 92.6%, sensitivity of 89.9%, and specificity of 95.2% (Figure 3d). Finally, the chloramphenicol phenotype model had a balanced accuracy of 88.8%, sensitivity of 78.3%, and specificity of 99.3% (Figure 3c). Inspection of the chloramphenicol-treated cells that were misclassified as untreated suggests that this model’s increased number of False Negative classifications was driven by cells that did not adopt the expected chloramphenicol-treated phenotype within the antibiotic treatment period, having multiple, diffuse nucleoid regions (Figure S4a), whereas False Positive classifications tended to have a central nucleoid region (Figure S4b). The balanced accuracies of >90% reported here are for single cells and therefore the cumulative accuracy of the classifier on a collection of cells is essentially 100%. Accuracy on a sample of cells is discussed further in the section on clinical isolates.

Ribosome phenotypes can be used to classify ciprofloxacin-resistant clinical *E. coli* isolates

Having demonstrated that antibiotic response phenotypes can be reliably induced and classified by a CNN, we moved to train a model to classify *E. coli* isolated from clinical samples as susceptible or resistant to ciprofloxacin using the ribosome phenotype. We called an isolate “resistant” if its minimum inhibitory concentration (MIC, the concentration required to inhibit overnight growth), was above the EUCAST breakpoint^9^. Isolates with MICs below the EUCAST breakpoint were called “susceptible”. We hypothesized that a CNN could learn to identify ribosome phenotypes associated with ciprofloxacin sensitivity or resistance, and that resistant cells would look similar to untreated cells^25^ following ciprofloxacin exposure.

To represent some of the variation present in pathogenic *E. coli*, we chose three susceptible strains (S1, S2, S3) and three resistant strains (R1, R2, R3), each with a different mutation in ciprofloxacin resistance-associated genes (Table 1). Each of the susceptible strains have a mutation in one of these genes, whereas the resistant strains all have three or more resistance-associated mutations. For example, strain R2 has two mutations in the GyrA, which encodes the A subunit of DNA gyrase; and three mutations in the genes that encode DNA topoisomerase IV (two mutations in parC, for the A subunit; and one in parE, for the B subunit). Each of these mutations has been associated with increased ciprofloxacin resistance in previous work^33–35^. Each isolate was treated with ciprofloxacin at a concentration previously determined to robustly induce phenotypic changes within 30 minutes (10 mg/L, 20X EUCAST breakpoint)^25^.

**Table 1.**
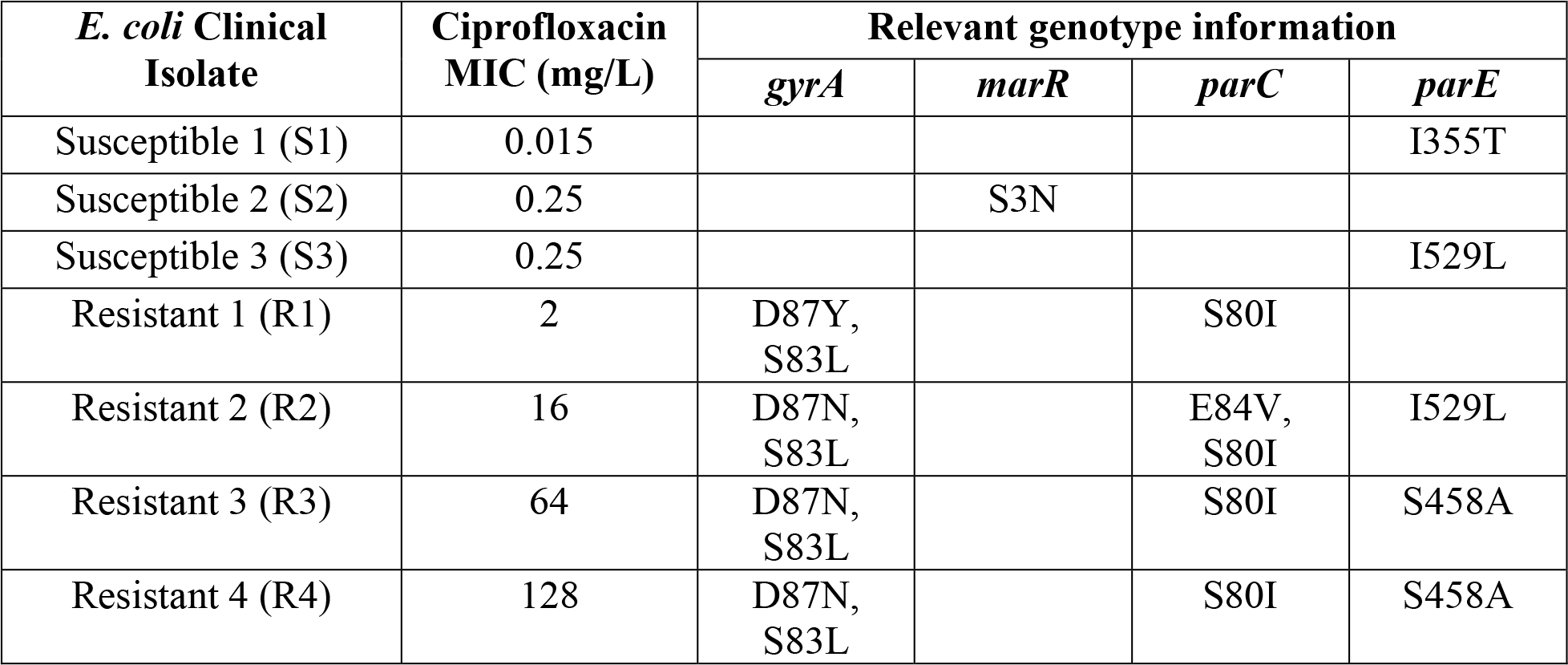
*E. coli* clinical isolates with their MICs and AMR genotypes. Each clinical isolate used in this project is listed with its MIC and relevant genotype information. All strains are Escherichia coli isolated from bloodstream infections in the United Kingdom, obtained and whole-genome sequenced for a previous study^44^. MICs were determined by broth microdilution. (See Methods: Bacterial strains and sample preparation for details of MIC and sequencing methods).

| *E. coli* Clinical Isolate | Ciprofloxacin MIC (mg/L) | Relevant genotype information |  |  |  |
| --- | --- | --- | --- | --- | --- |
|  |  | *gyrA* | *marR* | *parC* | *parE* |
| Susceptible 1 (S1) | 0.015 |  |  |  | I355T |
| Susceptible 2 (S2) | 0.25 |  | S3N |  |  |
| Susceptible 3 (S3) | 0.25 |  |  |  | I529L |
| Resistant 1 (R1) | 2 | D87Y, S83L |  | S80I |  |
| Resistant 2 (R2) | 16 | D87N, S83L |  | E84V, S80I | I529L |
| Resistant 3 (R3) | 64 | D87N, S83L |  | S80I | S458A |
| Resistant 4 (R4) | 128 | D87N, S83L |  | S80I | S458A |

Following ciprofloxacin treatment, all susceptible *E. coli* strains (S1, S2, & S3) demonstrated a ribosome phenotype similar to ciprofloxacin-treated *E. coli* MG1655, with a compact, central nucleoid region that we can detect indirectly because it results in low ribosome density in the central region (Figure 4a: S1, S2, S3). Some also showed an elongated morphology (Figure 4a: S1, S3). While some cells from the resistant strains resembled untreated *E. coli* MG1655, most cells from the resistant strains showed a different ciprofloxacin response, wherein the cells were elongated but retained diffuse nucleoid regions that spanned the cell length (Figure 4a: R1, R2, R3).

**Figure 4.**
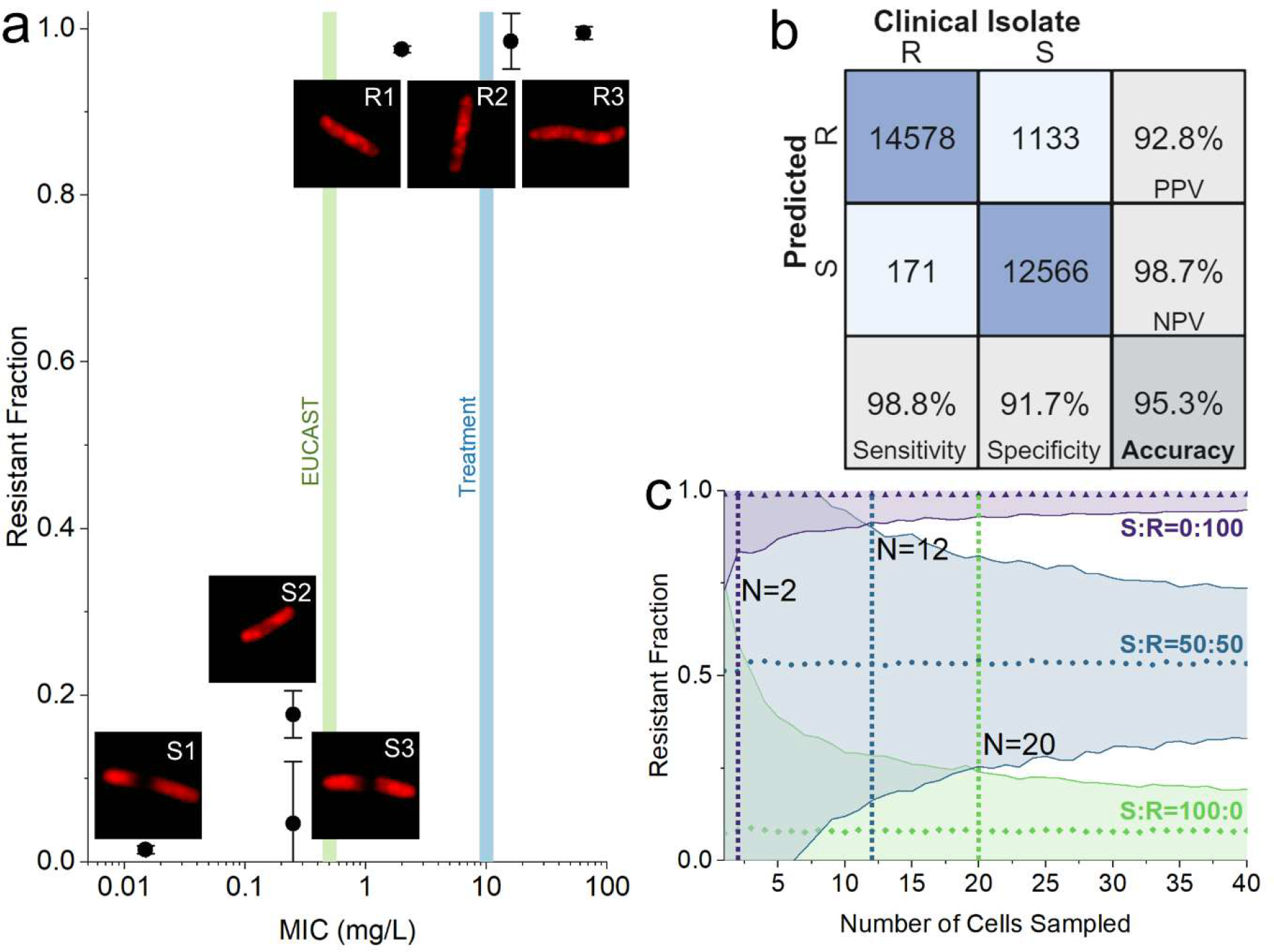
Susceptible (MIC < EUCAST breakpoint) and resistant (MIC > EUCAST breakpoint) *E. coli* isolates can be differentiated by the fraction of cells called resistant by the model. (a) The fraction of cells in the sample called resistant by the susceptible-resistant classifier (Resistant Fraction) is plotted against the MIC of the strain (mg/L) on a logarithmic scale, with error bars indicating the 95% confidence interval of the mean on two biological replicates. The test dataset is composed of holdout images, previously unseen by the classifier, from each clinical isolate. The EUCAST breakpoint (0.5 mg/L, green) and the treatment condition (10 mg/L, blue) are shown with shaded vertical lines. All strains with an MIC below the EUCAST breakpoint have a resistant fraction less than 0.2, whereas the fraction classified resistant is nearly 1.00 for the strains with an MIC above the EUCAST breakpoint. Representative, correctly classified images of ribosome phenotypes from each of the clinical isolates are shown for each point. Scale bars are not shown on the images because cells are resized to standardized 64x64 images before being passed to the CNN. (b) The confusion matrix for the ciprofloxacin-resistant and ciprofloxacin-susceptible classifier trained on 6 strains on a holdout, unseen dataset of 6 strains.The testing dataset is composed of 28,448 images from unseen biological replicates. See *Accuracy Metrics* for details on Accuracy, Sensitivity, Specificity, PPV, and NPV. (c) The number of cells necessary to classify a sample as coming from a population of susceptible or resistant bacteria. Simulated samples of different susceptible:resistant ratios (S:R) were transformed through the sensitivity and specificity of the susceptible-resistant classifier to determine the minimum number of cells necessary to differentiate them. Here, we plot the mean Resistant Fraction and 99% confidence interval after 1,000 trials with samples ranging from 1 to 40 cells sampled (N) for susceptible:resistant ratios of 0:100 (purple triangles), 50:50 (blue circles), and 100:0 (green diamonds). As the number of cells sampled increases, the confidence interval of the Resistant Fraction narrows. Susceptible samples can be differentiated from resistant samples with a sample of 2 cells (purple dotted line). A mixed sample can be differentiated from a resistant sample with 12 cells (blue dotted line) or from a susceptible sample with 20 cells (green dotted line). The confidence interval for resistant cells is narrower than that of susceptible cells because the classifier is more sensitive than it is specific.

In previous work, we have shown that a classifier trained on the nucleoid phenotypes of untreated and ciprofloxacin-treated *E. coli* MG1655 was able to classify susceptible and resistant clinical isolates accurately because resistant clinical isolates resembled untreated MG1655, while susceptible clinical isolates resembled ciprofloxacin-treated MG1655^25^. We applied this method to our ribosome images, using the CNN trained on ciprofloxacin-treated MG1655 to classify clinical isolates.

The MG1655 ciprofloxacin classifier had variable accuracy when applied to ciprofloxacin-treated clinical isolates (Figure S6a). The classifier recognized ciprofloxacin-susceptible phenotypes with high accuracy. For S1 and S3, it classified cells as ciprofloxacin-susceptible with 95.6±10.2% and 93.6±2.4% accuracy, respectively. The accuracy was slightly lower for S2 cells (84.6±5.4%), possibly because these cells are less likely to be elongated than ciprofloxacin-treated MG1655 (Mann-Whitney non-parametric hypothesis test p<0.05; Figure 4a, S2 isolate). For resistant isolates, the MG1655 classifier was less reliable. Compared to untreated MG1655, the ciprofloxacin-treated resistant isolates had similarly diffuse nucleoid regions but were elongated (Figure 4a: R1, R2, R3). The MG1655 classifier classified 90.6±2.8% of R2 cells as ciprofloxacin-resistant, but only 33.3±0.3% of R1 cells and 56.8±15.1% of R3 cells (Figure S6b). Representative cell images show that the R3 cells that were misclassified as susceptible had an elongated cell shape and a diffuse nucleoid (Figure S6a: R3), whereas R2 cells that were correctly classified as resistant had a shape and nucleoid phenotype more similar to the untreated MG1655 (Figure S6a, R2 isolate). The variability in accuracies for R1, R2, and R3 clearly show that the ribosome phenotypes resulting from ciprofloxacin treatment are too diverse to be reliably recognised by the MG1655 classifier, especially for resistant strains that develop an elongated shape with a diffuse nucleoid.

Therefore, we hypothesized that a CNN trained on images of ciprofloxacin-susceptible and resistant clinical *E. coli* isolates would be able to learn these variable responses and would perform better at the classification task. For this model, the training dataset was composed of 34,205 images of clinical *E. coli* treated with ciprofloxacin, which were segmented, zero-filled, and augmented as was done for the *E. coli* MG1655 model. We trained two six-strain models to check consistency on different biological replicates. For each of the six-strain models, two biological replicates were used for the training and validation datasets and one was used for a holdout test to assess the model’s accuracy on unseen data. In total, the testing dataset comprised 28,448 cells from three susceptible and three resistant clinical isolates (Figure 4b).

The susceptible-resistant CNN learned to identify phenotypes associated with ciprofloxacin-treated susceptible and resistant strains with a single-cell balanced accuracy of 95.3±8.3%; it displayed an accuracy of 98.8±2.3% in classifying resistant cells and an accuracy of 91.7±15.8% in classifying susceptible cells (Figure 4b). Because these high accuracies were on a per-cell basis, we were able to estimate the power of the model to classify an unknown population of *E. coli* as antibiotic-susceptible, antibiotic-resistant, or a mixture of the two. We simulated cell samples with 100% resistant cells, 100% susceptible cells, or a 50-50 mixture, using the sensitivity and specificity of our assay.

Given its high sensitivity (98.8%) and specificity (91.7%), our susceptible-resistant classifier has the power to differentiate a 100% resistant sample from a 100% susceptible sample with 99% confidence after sampling as few as 2 cells (Figure 4c). With a reasonable sample size of 10-100 bacteria isolated from a clinical specimen, the confidence level of our prediction would increase. In the case of a mixed infection or contaminated sample, we could differentiate a mixed sample from a resistant sample (12 cells) or a susceptible sample (20 cells) with the same level of confidence (Figure 4c). Stratifying the classifications by strain, we showed that our classification accuracy remained above 80% for all six strains (Figure S7). When examining the relationship between a given strain’s MIC and the fraction of cells classified as resistant, fewer than 20% of cells from strains with an MIC less than the EUCAST breakpoint were called resistant, whereas nearly 100% of cells from strains with an MIC above the EUCAST breakpoint were called resistant (Figure 4a). Compared to the MG1655 model, the susceptible-resistant model has similar or higher accuracy for all susceptible strains and is more accurate on all resistant strains (+64.1±.7% for R1, +7.9±6.1% for R2, +42.7±15.9% for R3) (Figure S8).

### Ribosome phenotypes can be used to classify unseen strains and antibiotic concentrations

To explore the generalizability of a CNN to previously unseen strains and antibiotic concentrations, another model was trained on just one susceptible (S2) and one resistant strain (R4). These strains showed characteristic ciprofloxacin-susceptible and ciprofloxacin-resistant phenotypes after 30 minutes treatment with 1X EUCAST ciprofloxacin (0.5 mg/L) (Figure 5a). This model was trained on a dataset of 2,888 cell images with an 80:20 training-validation split and tested on the same holdout dataset as the six-strain model, composed of unseen images from the six clinical isolates treated at 20X EUCAST (10 mg/L) for 30 minutes. The 1X EUCAST model was tested on each of the 3 biological replicates and performed with an average accuracy of 78.2±10.7% on cells from susceptible strains and an accuracy of 82.5±17.5% on cells from resistant strains (Figure 5b). Although lower in accuracy than the six-strain model, the two-strain model demonstrates an ability to reliably differentiate susceptible and resistant cells with relatively high accuracy while classifying images from never-before-seen strains treated at a different concentration of ciprofloxacin. This demonstrates the robustness of the ribosome phenotype classification method, so long as the model has seen sufficiently similar training data.

**Figure 5.**
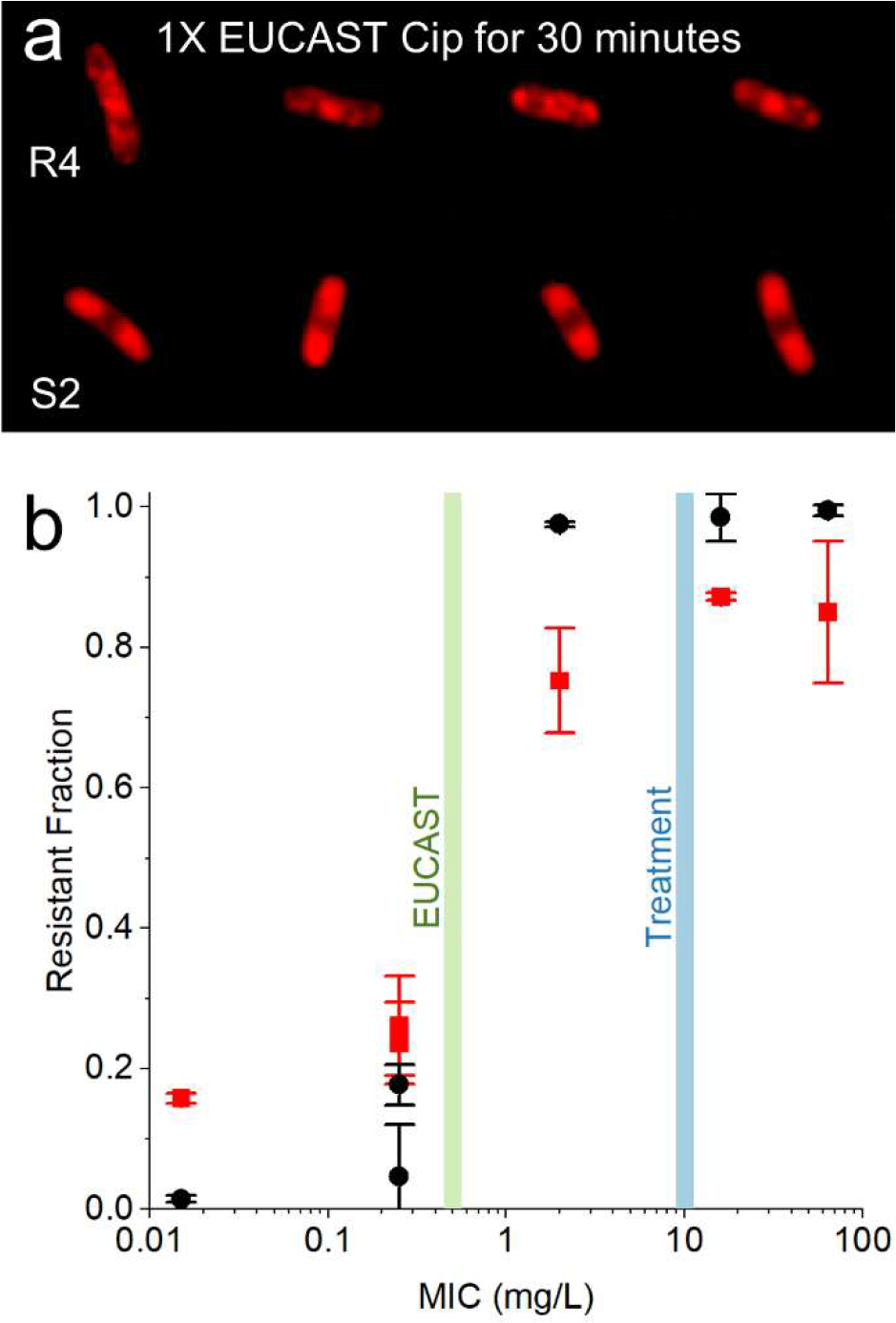
A CNN trained on isolates treated at 1X EUCAST maintains single-cell accuracy >75% on isolates treated at 20X EUCAST. (a) Representative, correctly classified images of the ribosome phenotypes of strains R4 and S2 treated at 1X EUCAST (0.5 mg/L) for 30 minutes. Scale bars are not shown because cells are resized to standardized 64x64 images before being passed to the CNN. (b) The susceptible-resistant classifier trained on 6 clinical isolates of *E. coli* treated with ciprofloxacin at 20X EUCAST (black circles) is compared to the classifier trained on 2 clinical isolates of *E. coli* (R4, S2) treated at 1X EUCAST (red squares). The error bars denote the 95% confidence interval of the mean accuracy. The 20X EUCAST model was tested on 28,448 holdout test images from 2 biological replicates of the 6 clinical *E. coli* isolates treated at 20X EUCAST ciprofloxacin for 30 minutes. The 1X EUCAST model was tested on 50,681 unseen images from the same 20X EUCAST dataset. For every isolate, the 20X EUCAST model is more likely to call resistant cells resistant and less likely to call susceptible cells resistant. However, the 1X EUCAST model maintains an accuracy of 78.2±10.7% on cells from susceptible strains and an accuracy of 82.5±17.5% on cells from resistant strains, despite being trained on images of cells treated at a different concentration and classifying previously unseen strains.

## Discussion

We have shown that ribosome-targeted FISH probes can be used to visualize intracellular antibiotic response phenotypes in *E. coli* that differ based on the mechanism of action of the antibiotic, and that these phenotypes can provide single-cell AST data with a single label. We demonstrated that distinct ribosome antibiotic response phenotypes exist for three clinically relevant antibiotics, and that these can be learned by a CNN with >90% accuracy. In clinical isolates of *E. coli*, we found that the antibiotic response can be more complicated, and it cannot be assumed that the resistant strain will always resemble the untreated phenotype. However, by using a model trained to identify the phenotypes of clinical isolates, we achieved an average single-cell classification accuracy of 95.3%. The ribosome phenotype classification method was also shown to extend to clinical strains not shown to the model in the training data and treated at a different antibiotic concentration. If deployed in a real-world AST, as the CNN models have access to training data from additional susceptible and resistant strains, the performance of the ribosome phenotype classifier would only be expected to improve.

In the context of a diagnostic test, our current single-cell accuracy means that with only 2 cells, we can differentiate a susceptible from a resistant sample with 99% confidence and can identify mixed infections with a sample of 20 bacterial cells. On a realistic scale of between 10 to 100 bacteria captured from a dilute sample such as blood or cerebrospinal fluid^36^, this level of accuracy could enable confident diagnosis even in less ideal imaging conditions. Compared to the previous deep-learning-based AST^25^ based on nucleoid and cell membrane staining, our ribosomal method achieves similar or greater accuracy while requiring only a single fluorescent label (92.1% vs. 91% on susceptible cells; 98.5% vs. 91% on resistant cells).

This previous deep-learning-based AST also showed that it could provide equivalent information to growth-based assays^25^ through the relationship between the proportion of cells classified as antibiotic-susceptible and the MIC of the strain. Here, using the ribosome phenotype, we also find a strong relationship between the MIC of the clinical isolate and its morphology after antibiotic treatment with ciprofloxacin. While the previous method used untreated lab-strain *E. coli* as a proxy for the resistant phenotype, we found that our MG1655-trained ribosome phenotype classifier had low accuracy when classifying resistant strains with a diffuse nucleoid but elongated cell shape. This could be because the MG1655 treated with ciprofloxacin is longer than untreated cells, and the ribosome phenotype classifier is classifying predominantly by cell length. Given the difference we found between clinical-strain and lab-strain phenotypes, we recommend that antibiotic response phenotypes are characterized in clinical isolates when possible, although lab strains can be used as a starting point.

The antibiotic response phenotypes of pathogenic *E. coli* are diverse, and there are many insights that could be learned from the 47,704 high-resolution single-cell images that were obtained for this study and are being made available for research (see Data Availability section). Generating these large, curated datasets of high-resolution bacterial images is time-intensive, because many high-throughput systems are optimised for eukaryotic cells, but they can be powerful in developing our understanding of bacterial antibiotic response. Beyond the heterogeneity in response within a single sample, we found that two isolates with the same MIC (0.25 mg/L) but different genotypes showed different morphologies and were classified as resistant at different rates (6% vs. 18%). These divergent responses form avenues for future research.

Compared to growth-based assays^14–16^, this method is limited to assessing the ribosome phenotype at a single timepoint because of the fixation and permeabilization steps necessary for FISH. The advantages of the ribosome-labelling FISH approach are the plethora of structural features that are amenable to deep-learning-based classification and the potential for simultaneous species ID. The combination of live-cell growth rate and fixed-cell phenotypic data could be even more powerful in assessing a cell’s antibiotic response.

Here, we demonstrate the accuracy of a ribosome phenotype AST on *E. coli* clinical isolates treated with ciprofloxacin. To have greater clinical utility, this method will need to be extended to other bacterial species and antibiotics. FISH probes targeting the ribosomal RNA have been used to identify a variety of Gram positive and negative species for clinical applications^37–40^. In the future, these probes could be combined with ribosome phenotyping. It is likely that the extension of this method from *E. coli* to other Gram-negative bacilli will be more straightforward, while the smaller size of cocci and lower permeability of Gram-positive bacteria may be a greater challenge. Similarly, we have shown that three antibiotics with intracellular targets (ciprofloxacin, gentamicin, chloramphenicol) cause characteristic ribosome phenotypes that can be identified by a CNN. We expect that this method can be extended to other antibiotics, so long as they reliably induce a visible change in the ribosome phenotype within the time scale of the test. As was shown for gentamicin (Figure S1), the benchmark treatment concentration may differ for each antibiotic. The length of antibiotic treatment may also need to be adjusted, especially for slow-growing species. Despite these challenges, when used in combination with bacterial genotyping, a single-cell imaging assay like this one could also be used to profile new resistance-associated mutations. This work serves as a guide for how deep learning can be used with fluorescence microscopy to learn intracellular phenotypes with high levels of accuracy, which can be applied to different species, and antibiotics, as well as to many biological, clinical and biotechnological applications.

In the context of ultra-rapid ASTs, although more conventional cytological profiling has advantages in interpretability^16,24^, the expected diversity of ribosome phenotypes in response to antibiotic treatment in different bacterial strains and species is one of the motivators for a CNN-based phenotypic AST, because a CNN can be expected to improve in performance when it is able to learn from additional, real-world data^41^.

By combining our single-cell ribosome-based assay with highly efficient microfluidic capture chips^36^, an AST could be performed on cells captured directly from the clinical specimen, eliminating the need for lengthy culture steps, and use multiplex FISH probes that bind to species-specific regions on the ribosomal RNA to report both the species ID and antibiotic susceptibility data.

## Methods

### Bacterial strains and sample preparation

*Escherichia coli* MG1655, a lab-adapted non-pathogenic K-12 derivative, was used as the reference strain for characterising antibiotic-susceptible ribosome phenotypes. Clinical strains were grown from stored blood culture isolates obtained for diagnostic and research purposes by the Microbiology Laboratory of the Oxford University Hospitals NHS Foundation Trust, Oxford, UK. All clinical isolates had been sequenced on the Illumina platform and AMR genotypes were assigned using the ResFinder^42^ database with Abricate v0.9.8^43^ (--min-id 95 –min-cov 95) as part of a previous study^44^ (Table 1).

The minimum inhibitory concentration (MIC) of each strain was tested in biological duplicate according to the broth microdilution method^45^ (Table 1, Table S1). The MIC was defined as the lowest antibiotic concentration inhibiting growth when the cultures were incubated overnight in Mueller-Hinton broth at 37℃.

Bacterial cultures were prepared in a shaking incubator at 37℃ in 5 mL lysogeny broth (MG1655) or Mueller-Hinton broth (clinical isolates) until reaching logarithmic growth, or OD600nm of 0.2. Then, antibiotics were added to reach the specified concentration (see EUCAST Breakout Points, Table S1) and the samples were returned to the incubator for the 30-minute treatment period. Samples were then fixed in 2% paraformaldehyde for 20 minutes. After fixation, the samples were centrifuged and the cell pellets were washed once with PBS, then re-centrifuged and re-suspended in 5 mL PBS before being split into 1 mL aliquots and permeabilised in 500 µL absolute ethanol before being stored at -20℃ until use.

Before imaging, the cells were centrifuged to remove the ethanol supernatant, washed with 500 µL PBS, and resuspended in hybridization buffer (20% v/v formamide, 0.9 M NaCl, 20 mM Tris pH 7.5, 0.01% SDS w/v). For labelling, 4’,6-diamidino-2-phenylindole (DAPI, 1 µg/mL) and 25 nM EUB338-Cyanine3 were added to the solution and the sample was incubated for 20 minutes. The ssDNA EUB338-Cyanine3 FISH probe has the sequence Cyanine3 – 5’ – gct gcc tcc cgt agg agt – 3’ (Sigma Aldrich). Following incubation, the samples were washed and resuspended in 150 µL PBS.

### Image acquisition

Samples were imaged on agarose pads prepared with 1.5% (w/v) high-purity agarose (Bio-Rad, catalogue number 1613101) in distilled water. Images were collected on the Nanoimager-S microscope (ONI, Oxford, UK) with a 100X oil-immersion objective in multi-acquisition mode. The DAPI stain was illuminated by a 405 nm laser in epifluorescence mode at a laser power of 5.1 kW/cm^2^ for an acquisition time of 20 ms. The Cyanine 3 fluorophore was illuminated with a 532 nm laser in epifluorescence mode at a laser power of 16.5 kW/cm^2^ for 20 ms.

### Image processing and segmentation

Each field of view was segmented using Napari-BacSeg^32^, a user-friendly bacterial analysis platform that allows microscopy images to be segmented using machine learning models, such as CellPose^31^ and OmniPose^46^. BacSeg can also be used to train custom CellPose or OmniPose models to improve segmentation performance and minimise the need to curate segmentations or fix segmentation errors. Within the software, the resulting segmentations can be easily curated and then exported in multiple formats to facilitate downstream analysis. Descriptive statistics of the segmented bacteria can also be computed and exported. The BacSeg Napari^47^ plugin can be installed from from the Napari Hub, the Python package manager PyPi, or GitHub (https://github.com/piedrro/napari-bacseg).

For our segmentations, custom CellPose^31^ models were trained on our 532 nm ribosome data for 100 epochs using the standard Napari-BacSeg hyperparameters to improve segmentation performance; these were then used for cell segmentation. Cells on the edge of the image, overlapping cells, vertical cells, or cells outside of the focal plane were removed from the final dataset during the segmentation and curation process.

### Cell phenotypes

From the curated segmentations, cell lengths, widths, and midlines were generated using the ColiCoords^48^ plugin within Napari-BacSeg^32^ with 10 midline vertices. For each cell, the intensity in each channel is normalised from 0 to 1 and the mean intensity is calculated for each channel for 100 bins along this midline. For a population of cells, the mean intensity and the standard deviation of the intensity is calculated for each of these 100 bins. This provides a mean intensity for each bin along the long axis of the cell. Because the cell lengths cannot be assumed to be normally distributed, hypothesis testing was conducted with the Mann-Whitney non-parametric test with a significance value of 0.05 in Origin Pro 2021.

### Neural network training

Images and segmentation maps were exported from Napari-BacSeg^32^ to create standardised 64x64 images of each cell zero-filled outside the segmentation boundary.

To account for different staining and illumination brightness, histogram normalisation was applied to each image. The images were randomly rotated, flipped, translated, sheared, and blurred with geometric and noise transforms from the Albumentations package^49^ before being loaded into the neural network.

The convolutional neural network was built with PyTorch^50^. Each model was run for 100 epochs with a batch size of 10, a learning rate of 0.001, and a training-validation split of 0.80/0.20. Optuna^51^ was used to optimise the hyperparameters for each model. The validation and testing datasets were balanced by class to reduce bias. Because the task was a single-label binary classification, DenseNet121^52^ with the Cross-Entropy loss function was used as the neural network structure. The Adam function^53^ was used for adaptive learning rates.

### Accuracy Metrics and Sample Size Simulation

Plots and phenotype statistics such as the Mann-Whitney non-parametric test were done using Origin Pro 2021. The Mann-Whitney non-parametric test was chosen because the phenotype measurements cannot be assumed to be normally distributed, as they are composed of cells at different stages of the cell cycle. The cell classification simulation to determine sample size was calculated using MATLAB R2022b.

1. Balanced Accuracy. All accuracies are reported with the 95% confidence interval, ± 2σ. Here we define resistant cells as “Positives” and susceptible cells as “Negatives.” For a one-class binary classification task,

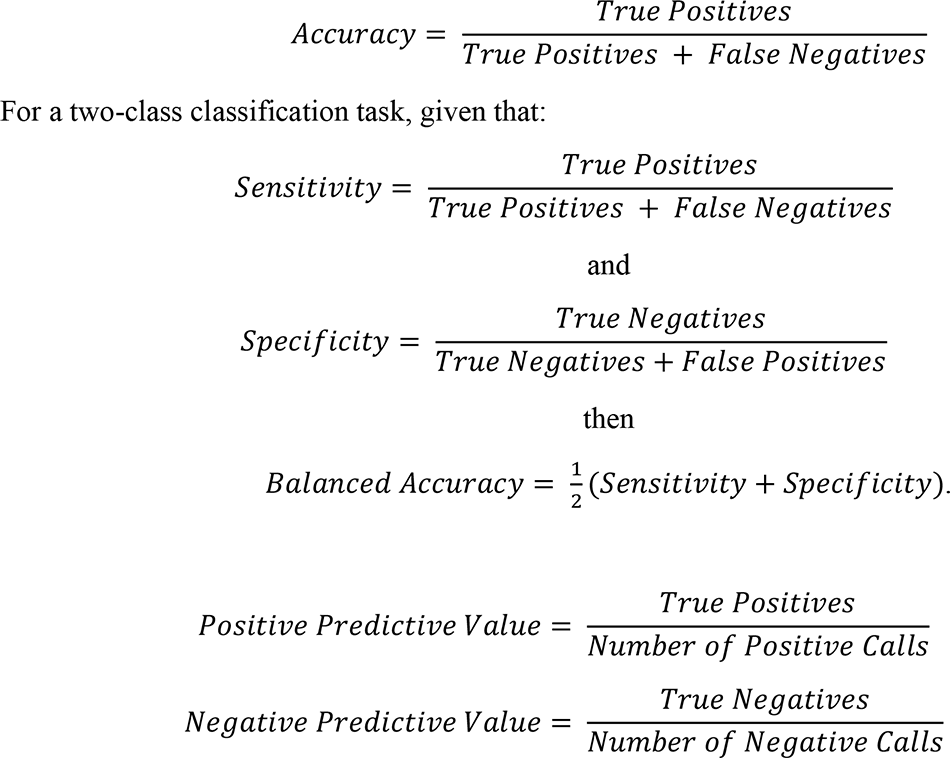

1. 2. Sample Size Simulation. Given a sample of cells of size N and a certain proportion of susceptible and resistant cells (100:0, 50:50, 0:100), we simulated the measured Resistant Fraction and 95% confidence interval. We simulated random samples of 10,000 susceptible and resistant cells at the defined proportions and transformed them into detected samples using the accuracy of our susceptible-resistant classifier. A resistant cell was detected as resistant 98.8% of the time (sensitivity) and a susceptible cell was detected as susceptible 91.7% of the time (specificity). Random populations of between 1-40 cells were sampled and the Resistant Fraction (resistant cells/total cells) was calculated. After 1,000 trials, the mean Resistant Fraction and 99% confidence interval (2.58σ) was plotted for each sample size. We defined the minimum sample size as the smallest sample for which the 99% confidence intervals did not overlap.

### Ethics

Ethical approval for the use of clinical isolates processed by the John Radcliffe Hospital microbiology laboratory in the development of diagnostic assays was granted by the UK’s Health Research Authority (London – Queen Square Research Ethics Committee [REC reference: 17/LO/1420]).

## Acknowledgements

This work was supported by the Oxford Martin School (by the establishment of the Oxford Martin School Programme on Antimicrobial Resistance Testing; to A.N.K., N.S., C.N., D.C. and M.A.), by Wellcome Trust grant 110164/Z/15/Z (to A.N.K.), by the Clarendon Fund Scholarships (to A.F.), and by UK Biotechnology and Biological Sciences Research Council grants BB/N018656/1 and BB/S008896/1 (to A.N.K.). The research was additionally supported by the National Institute for Health Research (NIHR) Health Protection Research Unit in Healthcare Associated Infections and Antimicrobial Resistance (NIHR200915) at the University of Oxford in partnership with United Kingdom Health Security Agency (UKHSA) and by the NIHR Oxford Biomedical Research Centre. The views expressed in this publication are those of the authors and not necessarily those of the NHS, NIHR, the Department of Health or Public Health England. Figures 1, 3 and 4 in this manuscript were created with the use of Biorender.com.

## Data availability

Code for model training is available at: https://github.com/KapanidisLab/ribosome_phenotype_classification.

Cell images and metadata are available at: https://zenodo.org/records/11656505.

## Conflict of interest

The work was carried out using a wide-field microscope from Oxford Nanoimaging, a company in which A.N.K. is a co-founder and shareholder. A.N.K. received no payment for this work from Oxford Nanoimaging, and Oxford Nanoimaging was not involved in any aspect of this work.

## Notes

### Author Declarations

Ethical approval for the use of clinical isolates processed by the John Radcliffe Hospital microbiology laboratory in the development of diagnostic assays was granted by the UK's Health Research Authority (London - Queen Square Research Ethics Committee [REC reference: 17/LO/1420]).

### Summary of Updates

Amended author list to include Hafez El Sayyed.

