## Supplementary Material for "Ribosome Phenotypes Enable Rapid Antibiotic Susceptibility Testing in *Escherichia coli*"

| | Antibiotic Minimum Inhibitory Concentration (mg/L) | | |
|---|---|---|---|
| | **Chloramphenicol** | **Gentamicin** | **Ciprofloxacin** |
| MG1655<br>K-12 Lab Strain | 3.75 | 0.125 | 0.016 |
| **EUCAST Breakout Point** | **>8** | **>2** | **>0.5** |

**Table S1. MG1655 MICs and EUCAST breakout points for chloramphenicol, gentamicin, and ciprofloxacin.** The minimum inhibitory concentration (mg/L) for *E. coli* MG1655 is shown for chloramphenicol, gentamicin, and ciprofloxacin. These concentrations were used to determine antibiotic treatment concentrations, e.g. 1X EUCAST = 8 mg/L chloramphenicol, 1X EUCAST = 0.5 mg/L ciprofloxacin, and 20X EUCAT = 40 mg/L gentamicin. MICs were determined by broth microdilution (see *Methods: Bacterial strains and sample preparation*).

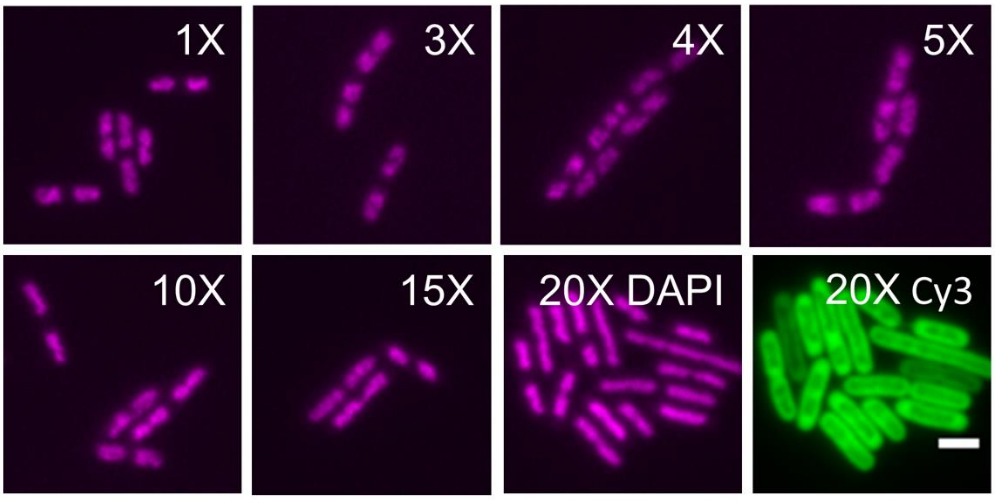


**Figure S1. Phenotypic changes associated with gentamicin treatment for 30 minutes at various concentrations from 1X to 20X EUCAST.** Representative images of MG1655 *E. coli* treated with gentamicin in LB media for 30 minutes, labelled with the gentamicin concentration in multiples of the EUCAST breakpoint (see Table 1). Images show the nucleoid (DAPI) unless otherwise labelled. Samples were labelled with DAPI and EUB338-Cy3 as described in the *Methods*. Scale bar, 2 μm.

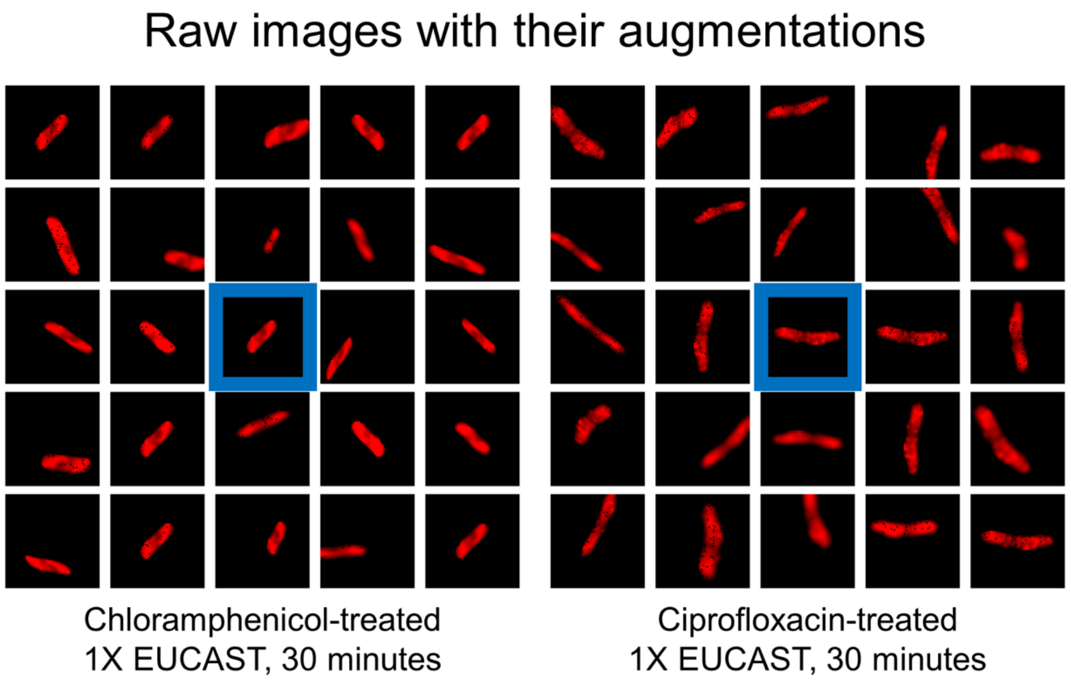


**Figure S2. Image augmentations.** Raw images (centre, highlighted in blue) and a selection of resulting augmented images are shown for representative chloramphenicol-treated and ciprofloxacin-treated cells. Scale bars are not shown because cells are resized to standardized 64x64 images before being passed to the CNN. Some of the augmentations can be seen, such as random rotations, flips, translations, shearing, blurring, and pixel dropout.

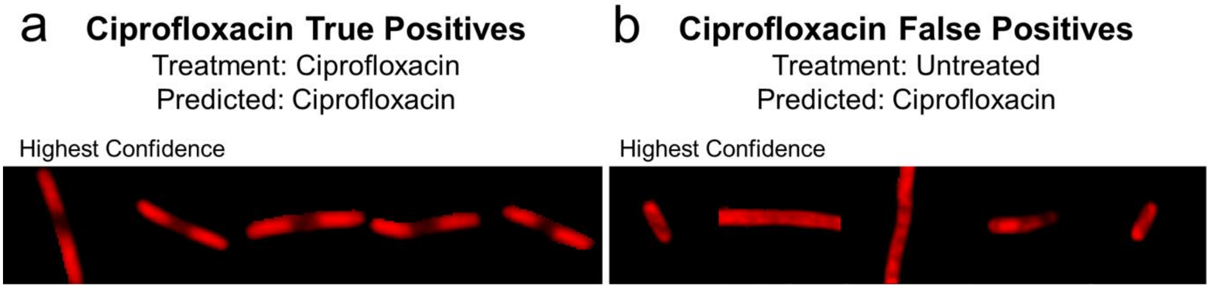


**Figure S3. Images of cells classified as ciprofloxacin-treated by the model.** Representative ribosome phenotypes of *E. coli* MG1655 are shown for (a) True Positives (ciprofloxacin-treated, predicted ciprofloxacin-treated) and (b) False Positives (untreated, predicted ciprofloxacin-treated) that were classified with the highest confidence by the model. Scale bars are not shown because cells are resized to standardized 64x64 images before being passed to the CNN. The True Positive images show an elongated phenotype and a central nucleoid region with lower ribosome intensity, especially in the images classified with the highest confidence. The False Positive images that were misclassified are elongated, or have a central nucleoid region, or both. This suggests that the ciprofloxacin model is using cell length and the existence of a central nucleoid to classify cells as ciprofloxacin-treated.

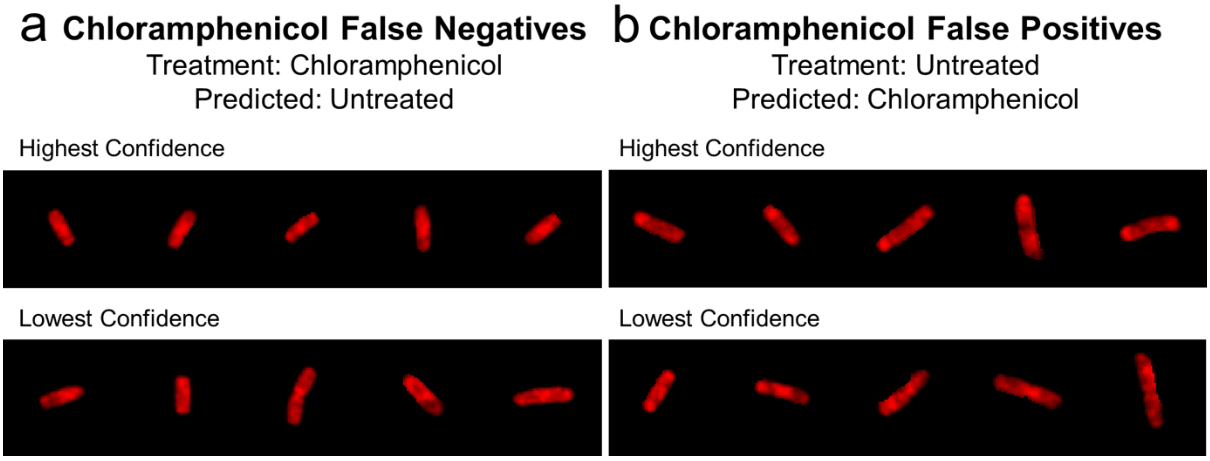


**Figure S4. Images of *E. coli* MG1655 incorrectly classified by the chloramphenicol model.** Example ribosome phenotypes of (a) False Negatives (chloramphenicol-treated, predicted untreated) and (b) False Positives (untreated, predicted chloramphenicol-treated). Scale bars are not shown because cells are resized to standardized 64x64 images before being passed to the CNN. The False Negative cells show multiple, diffuse nucleoid regions in chloramphenicol-treated cells, whereas the False Positives show central nucleoid regions in untreated cells.

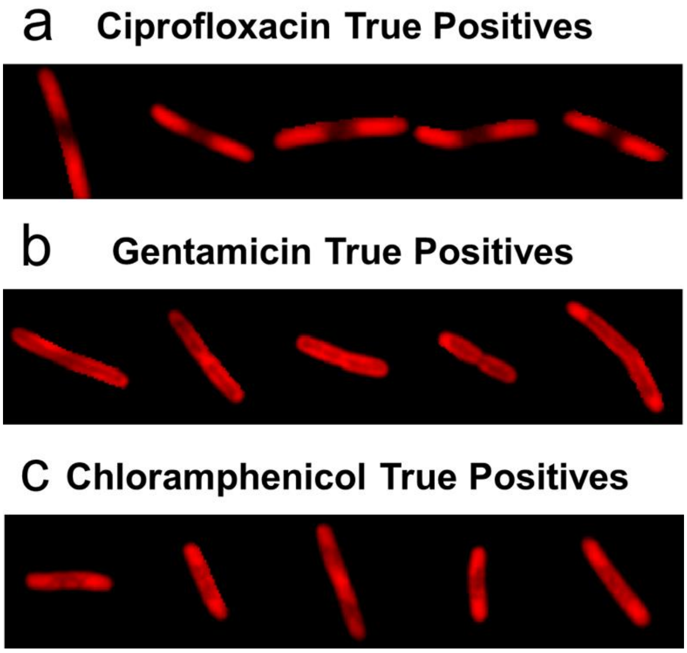


**Figure S5. Representative classifications for the ciprofloxacin, gentamicin, and chloramphenicol phenotype models.** Example ribosome phenotypes are shown for correctly classified *E. coli* MG1655 cells treated with ciprofloxacin (a), gentamicin (b), and chloramphenicol (c) as described in the *Methods*. The ciprofloxacin cells show elongated cells with a central, compact nucleoid region. The gentamicin cells show elongation and a nucleoid region that has compacted along the midline. The chloramphenicol cells show central nucleoid regions. Scale bars are not shown because cells are resized to standardized 64x64 images before being passed to the CNN.

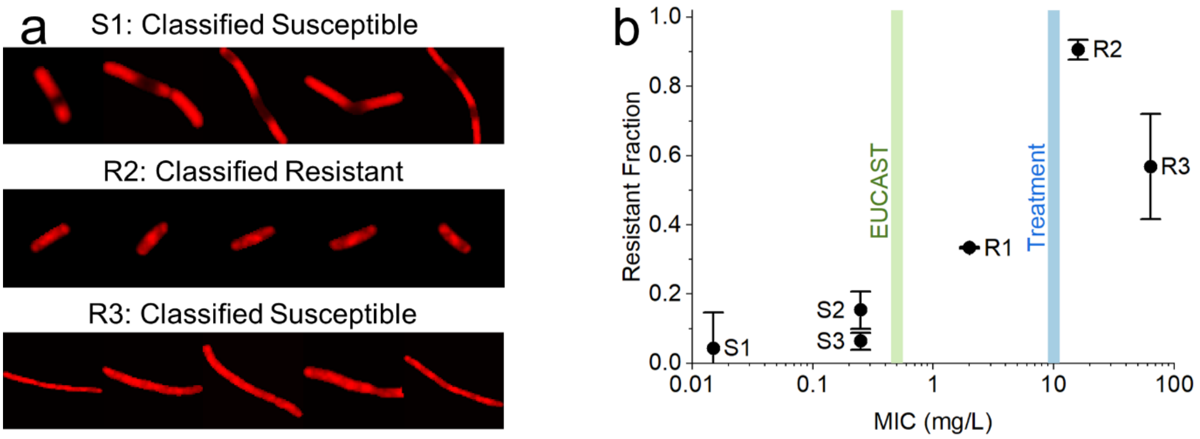


**Figure S6. Classifier trained on *E. coli* MG1655 applied to clinical isolates has variable accuracy.**

(a) Representative ribosome phenotypes from S1 cells correctly classified as susceptible, showing an elongated phenotype and compact nucleoid; R2 cells correctly classified as resistant, showing a phenotype more typical of untreated *E. coli* MG1655; and R3 cells incorrectly classified as susceptible, showing an elongated phenotype and diffuse nucleoid. Scale bars are not shown because cells are resized to standardized 64x64 images before being passed to the CNN.

(b) The fraction of cells in the sample called Resistant by the MG1655 ciprofloxacin classifier is plotted against the MIC of the strain (mg/L) on a logarithmic scale. Error bars represent the 95% confidence interval, equal to ± 2 standard deviations of the mean. The test dataset is composed of images from the six *E. coli* clinical isolates treated with 10 mg/L ciprofloxacin for 30 minutes. The EUCAST breakpoint (0.5 mg/L) and the treatment condition (10 mg/L) are shown with dashed vertical lines. All strains with an MIC below the EUCAST breakpoint have a resistant fraction less than 0.2, whereas there is no clear relationship between the MIC and the resistant fraction for the resistant strains.

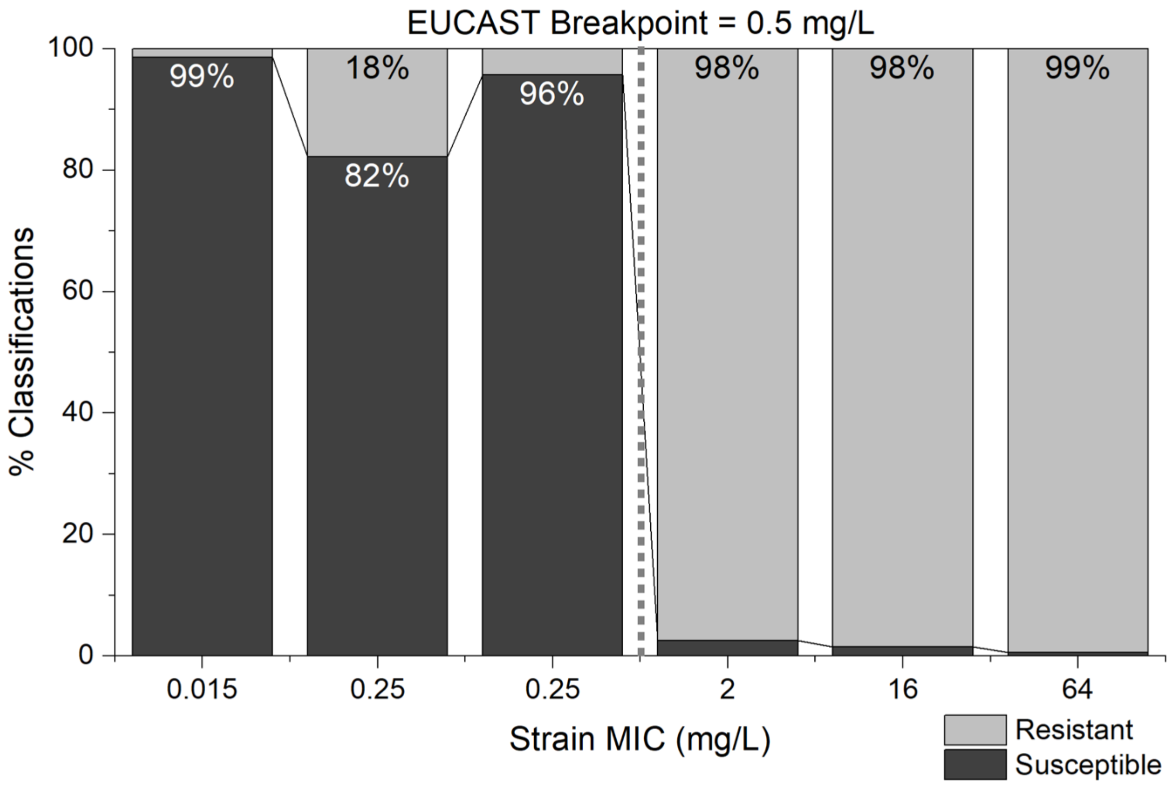


**Figure S7. The sensitive-resistant model can differentiate susceptible (MIC < 0.5 mg/L) and resistant (MIC > 0.5 mg/L) clinical isolates.** The percentage of cells in the test dataset classified as susceptible (dark grey) or resistant (light grey) is shown for each of the clinical isolates, ordered by the MIC of the strain. For strains with an MIC below the EUCAST breakpoint (0.5 mg/L), the % of susceptible classifications is greater than or equal to 80% (99%, 82%, 96%). For strains with an MIC above the EUCAST breakpoint, the % of resistant classifications approaches 100% (98%, 98%, 99%).

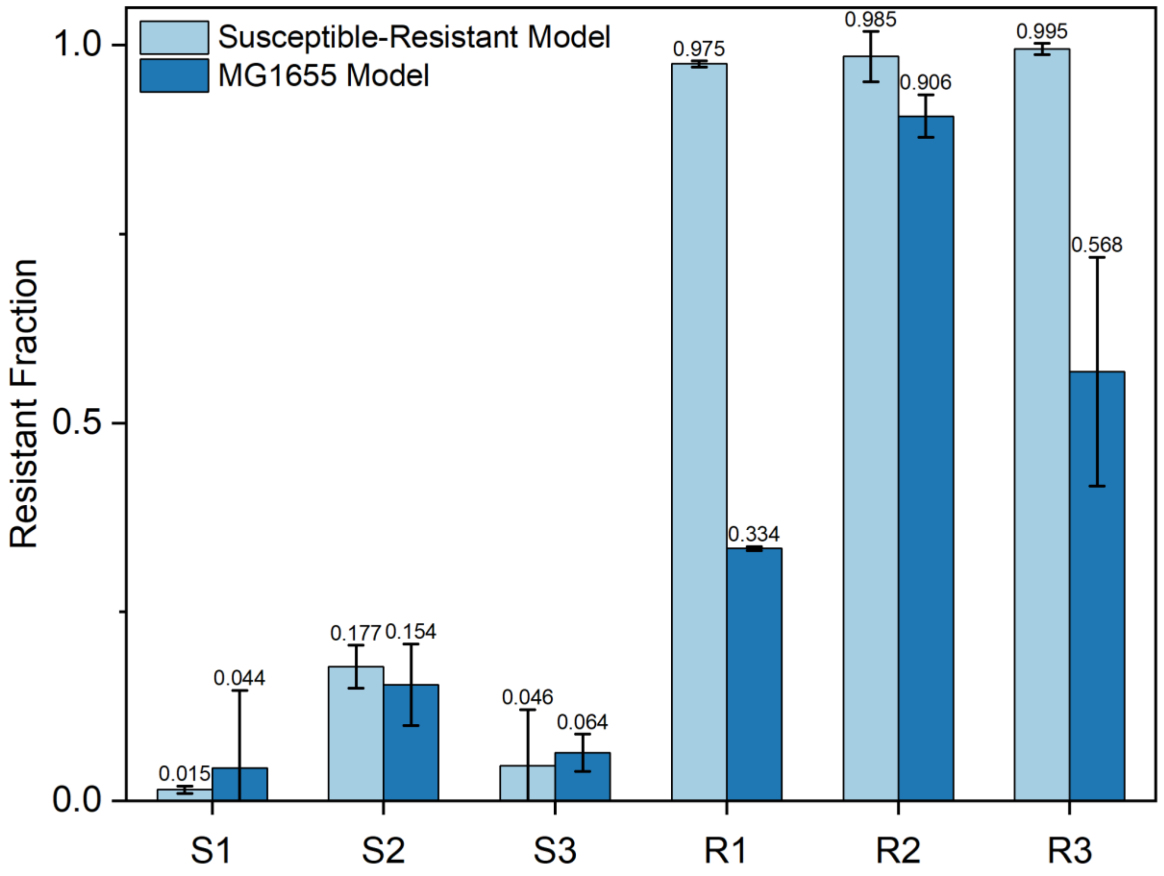


**Figure S8. The susceptible-resistant model based on clinical isolates is more accurate than the MG1655 model.** The fraction of cells classified as resistant (Resistant Fraction) is shown for each of the clinical strains (S1, S2, S3, R1, R2, R3), based on classifications by the susceptible-resistant model (light blue) or the MG1655 model (blue). The error bars denote the 95% confidence interval of the biological replicates. The test dataset is composed of images from the six *E. coli* clinical isolates treated with 10 mg/L ciprofloxacin (20X EUCAST) for 30 minutes. Compared to the MG1655 model, the susceptible-resistant model has similar or higher accuracy on all sensitive strains and is more accurate on all resistant strains.
